# Quantifying Uncertainty in ESBL *Enterobacterales* Transmission from Animals and Environment to Humans in Singapore: A Structured Expert Judgment Approach

**DOI:** 10.1101/2025.11.24.25340850

**Authors:** Yewei Xie, Nicholas Graves, Zhi Zhen Lim, Yin Mo, Isabelle Maya Srivastava, Yot Teerawattananon, Hwee Lin Wee, Alec Morton

## Abstract

**Objectives:** To estimate daily transmission rates of ESBL-producing Enterobacterales across hospital environment surface, animal, and community environmental settings in Singapore to address scarce empirical data that constrains One Health modelling and policy design.

**Methods:** A structured expert elicitation was conducted with a panel of eight experts: three for the clinical sector, three for the environmental sector, and two for the animal sector. Expert judgments were aggregated using both equal-weighting and Cooke’s Classical Model (performance-weighting) for hospital and companion-animal pathways, but only equal-weighting was used for the community-environment pathway.

**Results:** Under performance weighting, the daily transmission rate was 2.19 per person-day (95% Credible Interval [CrI] 0.16–10.47) for the hospital-surface pathway and 0.03 (95% CrI 0.00–0.11) for the companion-animal pathway. The corresponding equal-weighted estimates were 0.20 (95% CrI 0.01–2.77) and 2.13 (95% CrI 0.09–24.02), respectively. For the community-environment pathway (equal-weighted only), the rate was 0.81 (95% CrI 0.10–4.12).

**Conclusions:** These findings suggest that environmental and hospital pathways may represent important but uncertain contributors to ESBL-producing Enterobacterales spread in urban contexts, whereas animal-associated transmission appears limited. This study demonstrates that structured expert elicitation can provide systematic probabilistic priors for transmission and cost-effectiveness models, directly informing future resource allocation and the development of prevention and control strategies.

**Highlights:**

- We addressed the lack of empirical data on ESBL-producing *Enterobacterales* cross-setting transmission by using Structured Expert Elicitation to generate systematic, probabilistic priors for One Health models.
- Estimated daily transmission rates suggest that environmental and hospital pathways are uncertain but potentially significant contributors to ESBL spread, whereas animal-associated transmission appears limited in the urban context.
- These novel probabilistic priors can be directly embedded into cost-effectiveness models to inform future resource allocation and the development of data-driven antimicrobial resistance control strategies.

## Introduction

The global spread of antimicrobial resistance (AMR) is a major public health concern, with Extended-Spectrum Beta-Lactamase (ESBL) producing *Enterobacterales* significantly contributing to this growing problem (1,2). ESBL *Enterobacterales*, resistant to a wide range of beta-lactam antibiotics, cause difficult-to-treat infections, leading to increased morbidity, mortality, and healthcare costs (2–4). Combating AMR requires understanding the transmission dynamics of ESBL *Enterobacterales* across the complex network of human, animal, and environmental reservoirs, as emphasized by the One Health perspective (5).

ESBL *Enterobacterales* have been identified in various animal and environmental sources, including livestock, companion animals, wildlife, and water bodies (6–11), which can serve as potential sources of human exposure through direct contact, food consumption, or environmental contamination (12). However, quantifying the relative contribution of each reservoir to human infections and assessing transmission risks remain challenging due to the complicated One Health system, limited data availability, and variability in environmental conditions and human-animal interactions (13).

Addressing these knowledge gaps and informing evidence-based interventions requires innovative approaches that synthesize available information and provide reliable estimates of key transmission parameters. Structured expert elicitation, a well-established method that systematically collects and combines expert judgments, can quantify uncertain parameters in complex systems and bridge the gap between available data and information needed for decision-making in the context of AMR transmission (14,15).

This study aims to conduct a structured expert elicitation to gather expert judgments on critical parameters related to the transmission of ESBL *Enterobacterales* from animal and environmental reservoirs to humans. These parameters will inform mathematical models and risk assessments, providing insights into AMR transmission, guiding targeted interventions, and contributing to global efforts in combating AMR.

## Method

### Study design and expert selection

A structured expert elicitation was conducted to quantify key parameters related to the transmission of ESBL Enterobacterales from animal and environmental reservoirs to humans. Candidates were required to meet at least one of the following criteria: (1) hold a postgraduate degree (master’s or PhD) in a relevant field (e.g., microbiology, epidemiology, veterinary medicine, environmental science) with at least 3 years of post-degree professional experience in roles such as: clinical microbiologist, infectious diseases physician, antimicrobial resistance researcher, epidemiologist specializing in healthcare-associated infections, or veterinarian with antimicrobial stewardship experience; or (2) have a minimum of 5 years of professional experience in roles directly dealing with antimicrobial resistance issues in Singapore, such as clinical practice, research, policy-making, or regulatory oversight. Potential experts were identified through a combination of methods:

1. A targeted search of published literature on antimicrobial resistance in Singapore using databases such as PubMed and Scopus.
2. Review of staff directories and research profiles from relevant Singaporean institutions, including universities, hospitals, and government agencies.
3. Recommendations from key professional bodies and associations in Singapore related to infectious diseases, veterinary medicine, and environmental health.

The initial screening was based on publicly available information such as institutional profiles, publication records, and professional experience. Shortlisted candidates were contacted for further information and to confirm their willingness to participate. The expert elicitation comprised three separate assessments focusing on antibiotic resistance in Enterobacterales across animal, environmental, and clinical settings. We recruited a panel of eight experts in total: three for the clinical sector, three for the environmental sector, and two for the animal sector. Prior to the full elicitation, a pilot study was conducted with a small group of experts to test and refine the question format, ensure clarity, and identify any potential issues with the elicitation process. This piloting phase helped optimize the effectiveness of the main elicitation and improve the quality of the data collected.

### Elicitation parameters and procedure

Experts expressed their uncertainty about unknown quantities by providing specified quantiles from their subjective probability distributions. These assessments were provided for two types of questions: variables of interest and calibration questions (also known as “seed questions”).

1. Variables of Interest

These are the primary focus of the elicitation and are questions that cannot be adequately addressed with existing data or models, necessitating expert judgment. The primary variable of interest is 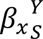, the per-capita, per-day hazard at which susceptible individuals acquire colonisation after contact with a antibiotic resistant reservoir in the environment (Y=E) or colonizing an animal (Y=A), whether in the hospital setting (X=H) or the community setting (X=C). We drew from the literature on spillover force of infection terms to assume the process of colonization for susceptible individuals in contact with a contaminated environment or contaminated animals (16). We assumed that the transmission coefficient can be calculated as the product of the prevalence of resistant bacteria in the reservoir (the environment or animals), the reservoir-human contact rate, and the probability of infection given contact (or transmissibility).

- We parameterize the reservoir prevalence as ρ_A_ for animals and ρ_E_ for the environment.
- We parameterize the reservoir-human contact rate as c_A_ for animals and c_E_ for the environment.
- We parameterize the probability of successful transmission as φ_A_ for animals and φ_E_ for the environment.

Hence, we have the following parameters (table 1):

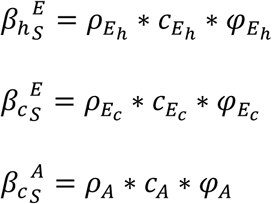

**Table 1:** Estimated parameters and daily transmission rates for ESBL-producing Enterobacterales transmission by sector.

| **Settings** | **Parameter** | **Equal-weighted Median (CrI)** | **Performance-weighted Median (CrI)** |
| --- | --- | --- | --- |
| **Hospital Surface environment** | Daily contacts with hospital surfaces | 73 (11–1392) | 699 (14–1467) |
|  | Prevalence on hospital surfaces (%) | 13.00 (2.00–29.00) | 10.00 (1.00–24.00) |
|  | Per-contact transmission probability (%) | 1.00 (0.00–5.00) | 1.40 (0.02–5.00) |
|  | Daily transmission rate | 0.20 (0.01–2.77) | 2.19 (0.16–10.47) |
| **Companion Animals** | Daily human–animal contacts | 34 (2–183) | 25 (1–40) |
|  | Prevalence in animals (%) | 17.00 (0.00–72.00) | 4.00 (0.00–10.00) |
|  | Per-contact transmission probability (%) | 1.00 (0.00–48.00) | 3.00 (0.00–5.00) |
|  | Daily transmission rate | 2.13 (0.09–24.02) | 0.03 (0.00–0.11) |
| **Community Environment** | Daily contacts with environmental surfaces | 40 (7–97) | NA |
|  | Prevalence in environment (%) | 18.00 (3.00–36.00) | NA |
|  | Per-contact transmission probability (%) | 11.70 (3.00–23.00) | NA |
|  | Daily transmission rate | 0.81 (0.10–4.12) | NA |
| Values are presented as medians with corresponding 95% credible intervals (CrI). NA = not applicable. |  |  |  |

2. Calibration Questions

These are items closely related to the variables of interest, but for which the true values are known to the study team, either at the time of the expert interview or later during the study period. Calibration questions enable empirical validation of the experts’ hypotheses. Due to data limitations, calibration questions were developed only for the human and animal sectors. The calibration questions were informed by an unpublished systematic review conducted by the study team, which synthesized estimates of the prevalence of antibiotic resistance among Enterobacterales for human and animal sectors in Singapore from 2013 to 2023 (17). Seed question data were drawn from the review’s appendix, which was unavailable to experts during elicitation as the manuscript was under peer review. Detailed descriptions of the calibration questions are provided in the appendix.

Table 1. Parameters selected for expert elicitation.

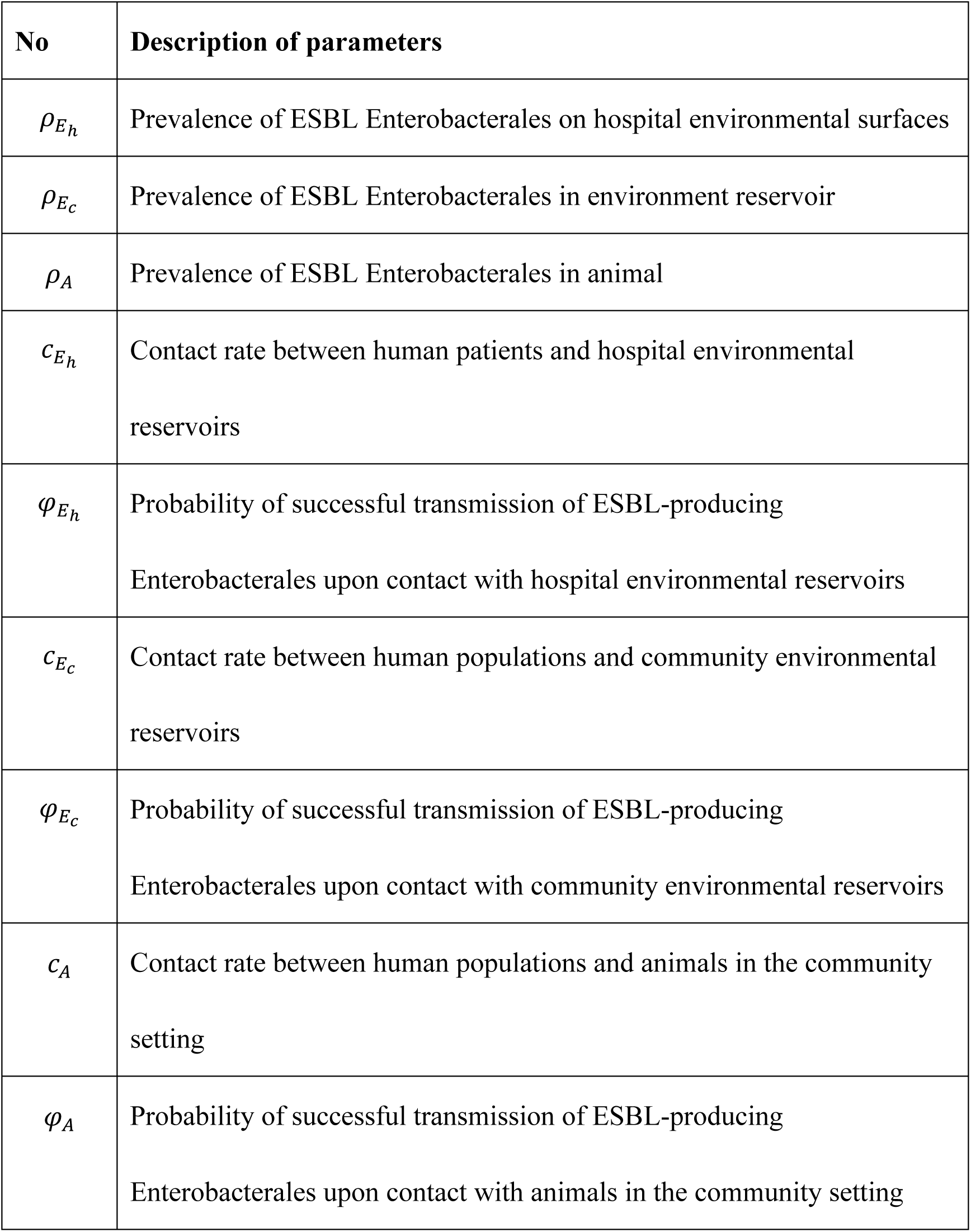

Prior to the elicitation, experts were informed that the goal was to quantify uncertainty rather than obtain precise estimates, and discussions were held to address cognitive biases and ensure reliable parameter estimation. One-on-one interviews were conducted with each expert, lasting between 30 and 60 minutes by Zoom, following a standardized protocol to ensure consistency. During the elicitation, experts were asked to quantify their beliefs on the transmission coefficients and their component parameters. Probability distributions were shown to the experts to confirm that they accurately represent their judgments.

### Data analysis and aggregation

Expert judgments were aggregated separately for the hospital surface environment, animal, and environmental settings. Across all sectors, an initial aggregation was conducted using an equal-weighted model, where each expert’s judgments contributed equally to the pooled distributions for each parameter of interest.

For the hospital surface environment and animal settings, we applied Cooke’s Classical Model to aggregate expert judgments on the variables of interest. The Classical Model is a structured expert judgment method that assigns weights to experts based on their performance in calibration questions and the informativeness of their judgments (18,19). The aggregated probability distribution for each parameter *θ* were calculated using the following formula:

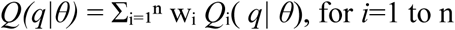

where n is the number of experts, *Qᵢ(q|θ)* is expert i’s estimate for quantile q of parameter *θ*, *Q(q|θ)* is the aggregated quantile estimate, and *wᵢ* are the Classical Model weights based on experts’ statistical accuracy and information scores. Continuous probability distributions were then fitted to these aggregated quantiles.

Experts answered ten calibration questions in the hospital surface environment and animal settings, enabling evaluation of statistical accuracy (calibration) and informativeness according to Cooke’s Classical Model. Performance-based weights were derived through global optimization, combining calibration and information scores to form a weighted aggregation referred to as the decision maker. Aggregated distributions were calculated as weighted linear combinations of individual expert estimates. In the environmental sector, where no suitable empirical data for calibration could be identified, expert judgments were aggregated using equal weights. All analyses were conducted using Excalibur software.

Daily transmission rates (β) were calculated as the product of three uncertain quantities: daily contacts, pathogen prevalence, and per-contact transmission probability. We employed Monte Carlo simulation with 1000 iterations, sampling from fitted distributions for each component parameter to calculate transmission rates for each iteration. The 5th and 95th percentiles of the resulting distributions formed the 90% credible intervals. All analyses were conducted in R.

### Ethics considerations

The IRB exemption of this study was acquired from Saw Swee Hock School of Public Health, National University of Singapore (NUS-IRB-2024-824).

## Results

### Hospital Surface Environment

The equal-weighted aggregation yielded median estimates of 73 contacts per day (90% Credible Interval [CrI] 11–1392) for human–hospital environmental surface contacts, 13.00% (90% CrI 2.00–29.00) for the prevalence of ESBL-producing *Enterobacterales* on hospital surfaces, and 1.00% (90% CrI 0.00–5.00) for the per-contact transmission probability. The daily transmission rate per patient for human-hospital surface environment contact was 0.2 (90% CrI 0.01–2.77).

Expert calibration using Cooke’s Classical Model identified Expert 1 as having the highest combined score (0.21), followed by Expert 3 (0.12) and Expert 2 (0.01) (Table 2). Performance-weighted aggregation produced slightly higher median estimates of 699 contacts per day (90% CrI 14–1467), 10.00% (90% CrI 1.00–24.00) for prevalence, and 1.40% (90% CrI 0.02–5.00) for the per-contact transmission probability, resulting in a daily transmission rate of 2.19 (90% CrI 0.16–10.47) (Table 2).

**Table 2.** Calibration Results for Experts in Human and Animal Sectors.

| Sector | Expert ID | Statistical Accuracy | Information Score |  | Combined Score | Normalized Weight (without DM) | Normalized Weight (with DM) |
| --- | --- | --- | --- | --- | --- | --- | --- |
|  |  |  | All Variables | Calibration Variables |  |  |  |
| **Hospital Surface environment** | Expert 1 | 0.44 | 0.47 | 0.44 | 0.21 | 0.63 | 0.34 |
|  | Expert 2 | 0.02 | 0.63 | 0.51 | 0.01 | 0.00 | 0.00 |
|  | Expert 3 | 0.34 | 0.37 | 0.34 | 0.12 | 0.37 | 0.20 |
|  | Performance-weighted DM | 0.96 | 0.27 | 0.27 | 0.26 | NA | 0.46 |
|  | Equal-weighted DM | 0.26 | 0.17 | 0.15 | 0.04 | NA | NA |
| **Companion Animals** | Expert 1 | 0.00 | 0.32 | 0.29 | 0.00 | 0.00 | 0.00 |
|  | Expert 2 | 0.00 | 1.12 | 0.97 | 0.00 | 1.00 | 0.50 |
|  | Performance-weighted DM | 0.00 | 1.12 | 0.97 | 0.00 | NA | 0.50 |
|  | Equal-weighted DM | 0.01 | 0.22 | 0.18 | 0.00 | NA | NA |
| DM: Decision maker. |  |  |  |  |  |  |  |

### Companion Animals

In the animal sector, equal-weighted aggregation produced median estimates of 34 human–animal contacts per day (90% CrI 2–183), 17.00% (90% CrI 0.00–72.00) for the prevalence of ESBL-producing *Enterobacterales* in animals, and 1.00% (90% CrI 0.00–48.00) for the per-contact transmission probability (Table 1). The daily transmission rate per pet owner was 2.13 (90% CrI 0.09–24.02).

Cooke’s method identified Expert 2 as having superior calibration performance (combined score 0.01) compared to Expert 1 (0.00) (Table 2). Performance-weighted aggregation produced median estimates of 25 human–animal contacts per day (90% CrI 1–40), 4.00% (90% CrI 0.00–10.00) for prevalence, and 3.00% (90% CrI 0.00–5.00) for the per-contact transmission probability, resulting in a daily transmission rate of 0.03 (90% CrI 0.00–0.11) (Table 1).

### Community Environment

For the community environmental setting, due to the lack of calibration questions, only equal-weighted aggregation was applied (Table 1). The median estimates were 40 contacts per day (90% CrI 7–97) for human–environmental surface contacts, 18.00% (90% CrI 3.00–36.00) for the prevalence of resistant organisms in the environment, and 11.70% (90% CrI 3.00–23.00) for the per-contact transmission probability. The daily transmission rate per person for general human-community environmental surface contact was 0.81 (90% CrI 0.10–4.12).

## Discussion

In this study, we employed structured expert elicitation to estimate key parameters influencing the transmission of ESBL-producing *Enterobacterales* across different urban settings. We applied this method to generate plausible ranges for contact rates, prevalence of ESBL-producing *Enterobacterales*, and transmission probabilities, providing informative priors for future modeling in settings where empirical data are limited. This study provided a structured, expert-informed foundation to support parameterization of future One Health models in the absence of reliable empirical data.

Our study estimated the daily transmission rate of ESBL-producing *Enterobacterales* from hospital environmental surfaces at 0.2 to 2.19, depending on the aggregation method. This estimate suggests that contact with contaminated surfaces is a plausible pathway for patient colonization, providing a quantitative starting point for understanding the contribution of environmental reservoirs to healthcare-associated transmission. These findings align with observational studies documenting persistent contamination of hospital sinks, drains, and high-touch surfaces with ESBL-producing organisms and implicating these sites in cross-transmission events (20–22). The broad CrI underscores variability in cleaning efficacy and contact heterogeneity, emphasizing the need for context-specific interventions like enhanced environmental monitoring.

Relatively low daily transmission rate of ESBL-producing *Enterobacterales* from animal sources were identified compared to other estimations in our study. This finding is consistent with population-based modelling studies attributing a relatively small proportion of community-acquired resistant *Enterobacterales* carriage to animal reservoirs in countries with high levels of urbanization and low direct animal contact (11,23). The estimate may reflect Singapore’s strong veterinary antimicrobial regulations. Future research incorporating molecular epidemiology and One Health surveillance would be valuable to confirm the limited zoonotic contribution and to monitor potential shifts in animal-associated transmission under changing agricultural or environmental conditions.

In contrast, our study estimated a higher daily transmission rate of ESBL-producing *Enterobacterales* from community environmental reservoirs, approaching levels estimated for hospital environmental surfaces and suggesting a potentially underrecognized role of environmental exposure in community transmission. This estimate diverges from prior modelling studies that attributed a smaller contribution to environmental pathways (11), raising the possibility that Singapore’s dense urban environment, high-contact public spaces, and wastewater infrastructure may elevate environmental exposure risks. Supporting this hypothesis, environmental surveillance studies have documented the presence of ESBL-producing *Enterobacterales* in urban waterways and environmental samples (9,10,24). These findings highlight the need for targeted environmental surveillance and exposure assessment in Singapore to better quantify environmental contamination sources and inform public health interventions within a One Health framework.

Moreover, the wide credible intervals across all transmission pathways reflect substantial uncertainty in current parameter estimates, highlighting critical knowledge gaps. While additional empirical studies could reduce these uncertainties, limited research resources necessitate strategic prioritization of which parameters warrant immediate investigation. For future studies, value of information analyses are needed to quantify the expected benefits of reducing specific parameter uncertainties to prioritize future research and resource allocation (25,26).

The transmission rates estimated in this study have direct implications for One Health modelling and public health action. By providing probabilistic priors to replace assumed or fixed values, our work enables models to explore uncertainty more fully and to test intervention effects under both median and extreme scenarios. The wide credible intervals for certain parameters also provide a systematic basis for prioritising surveillance, indicating where new empirical data would most effectively reduce policy-relevant uncertainty. Ultimately, these estimates allow models to compare the relative importance of human, animal, and environmental transmission, thereby supporting the strategic allocation of resources toward the most influential pathways.

Our study is subject to several limitations. Although structured elicitation improves on informal judgment, our estimates are necessarily influenced by the experiences and disciplinary backgrounds of the expert panel. Furthermore, calibration could only be applied to questions regarding the human and animal sectors. The absence of robust data for environmental parameters prevented similar validation, which reduced their robustness and widened their credible intervals. The estimates are also most applicable to densely urbanised settings and might not generalise to rural or agrarian contexts where exposure dynamics differ substantially. These limitations highlight a clear agenda for future research. Value of information analyses should guide investment decisions by quantifying the expected benefits of reducing specific parameter uncertainties. Once priorities are established through such analyses, refining these parameters will require longitudinal sampling in hospitals, environmental monitoring linked to genomic attribution, and dedicated studies of human–animal contact patterns to advance One Health transmission models.

## Conclusions

This study addresses a critical barrier in One Health antimicrobial resistance research by providing the first systematic, expert-derived probabilistic estimates for inter-sectoral ESBL-producing *Enterobacterales* transmission. These quantitative priors are essential for developing more robust dynamic models that can accurately explore uncertainty and evaluate cross-sectoral interventions. By translating expert knowledge into actionable data, our work provides a foundational and transferable framework for guiding surveillance, optimising resource allocation, and ultimately designing more effective strategies to combat antimicrobial resistance in complex urban settings worldwide.

## Supporting information

Seed question data were drawn from the review's appendix, which was unavailable to experts during elicitation as the manuscript was under peer review.

## Contributors

YX, NG, YM, and AM conceived and designed the modelling study. YX and YM curated the data and conducted the formal analysis. ZL, YT, IS, and HW provided methodological input and contributed to interpretation. YX drafted the manuscript. All authors revised the manuscript critically for important intellectual content, had full access to all data, approved the final version, and accept responsibility for the integrity of the work.

## Data sharing

Requests for data by researchers with proposed use of the data can be made to the corresponding author with specific data needs, analysis plans, and dissemination plans.

## Data Availability

All data produced in the present study are available upon reasonable request to the authors.

## Acknowledgments

This study was supported by the partners under Singapore’s One Health National Strategic Action Plan: the Ministry of Health, National Environment Agency, National Parks Board, PUB (Singapore’s National Water Agency), and Singapore Food Agency (Grant No. OHARP-2022-003); and by the SingHealth Duke-NUS Global Health Institute (Grant No. SDGHI_PHDRG_FY2024_0004-01). The authors are entirely responsible for the content of this manuscript, and the views and opinions expressed in this publication are solely those of the authors and do not necessarily reflect those of the funders.

