## Supplementary material for "Quantifying Uncertainty in ESBL *Enterobacterales* Transmission from Animals and Environment to Humans in Singapore: A Structured Expert Judgment Approach": Seed question data were drawn from the review's appendix, which was unavailable to experts during elicitation as the manuscript was under peer review.

**Appendix 1: Elicitation protocol**

**Introduction**

Structured expert judgment is a recognized tool in risk analysis, used to address data shortfalls, quantify uncertainty, and build consensus. It has been applied in studies sponsored by entities such as the European Union, World Health Organization, and the Robert Wood Johnson Foundation. This method is particularly valuable in contexts where repeated experimentation or rigorous data collection is not feasible. This study aims to conduct a structured expert elicitation to gather expert judgments on critical parameters related to the transmission of ESBL Enterobacterales from animal and environmental reservoirs to humans. These parameters will inform mathematical models and risk assessments, providing insights into AMR transmission, guiding targeted interventions, and contributing to global efforts in combating AMR.

**Expert names**

Expert names are preserved to enable competent peer review but are not associated with responses in any open documentation. Expert reasoning is captured during the elicitation and becomes part of the published record. Both the quantitative estimates and the qualitative reasoning from the experts are anonymous in any published report or article.

**Elicitation Format**

The elicitation consists of specifying percentiles of uncertain quantities, as illustrated below.

You are presented with an uncertainty quantity:

| In Singapore, what percent of the sampled Enterobacterales isolates from human sector are ESBL-producing? | | | | |
| --- | --- | --- | --- | --- |
| 5% | 25% | 50% | 75% | 95% |

Although the true value to these example questions based on the data is unknown, presumably, you don’t know the number off the top of your head, so the number you have in mind is uncertain.

You are asked to provide numbers that quantify your uncertainty by specifying these five percentiles:

The 5th percentile is the number for which you think there is a 5% chance the true value is below and a 95% chance the true value is above.

The 25th percentile is the number for which you think there is a 25% chance the true value is below and a 75% chance the true value is above.

The 50th percentile is the number for which you think there is a 50% chance the true value is below and a 50% chance the true value is above.

The 75th percentile is the number for which you think there is a 75% chance the true value is below and a 25% chance the true value is above.

The 95th percentile is the number for which you think there is a 95% chance the true value is below and a 5% chance the true value is above.

The values you provide for a question must always be increasing, such that:

5th percentile < 25th percentile < 50th percentile < 75th percentile < 95th percentile

**Example Responses**

Suppose you answer as shown below:

| In Southeast Asia, what percent of the sampled Enterobacterales isolates from human sector are ESBL-producing? | | | | |
| --- | --- | --- | --- | --- |
| 10 | 18 | 20 | 35 | 60 |
| 5% | 25% | 50% | 75% | 95% |

This means you believe the true value is equally likely to be above or below 20%. You believe there is a 50% chance the true percent of isolates that are ESBL-producing falls between 18% and 35%. You believe there is a 90% chance the true value falls between 10% and 60%. The actual value for this question is unknown, but if it falls within your specified 90% confidence band, you would not be surprised by the answer. However, you would have been surprised to hear the true value were 5% or 75%.

**Training**

Below are a few practice questions to familiarize you with the format and performance concepts.

The following questions are based on data for Southeast Asia in recent years.

| T1. In Southeast Asia, what percent of the sampled Enterobacterales isolates from human sector are resistant to carbapenems? | | | | |
| --- | --- | --- | --- | --- |
| 5% | 25% | 50% | 75% | 95% |

| T2. In Southeast Asia, what percent of the sampled Enterobacterales isolates from human sector are resistant to third generation cephalosporins? | | | | |
| --- | --- | --- | --- | --- |
| 5% | 25% | 50% | 75% | 95% |

**Elicitation process**

A. **Calibration questions**

We are seeking your expertise on the prevalence of ESBL-producing Enterobacterales and related antimicrobial resistance patterns in Singapore, encompassing data from both hospital and community settings. The dataset for the calibration questions is derived from an unpublished systematic review and meta-analysis of literature published between January 2013 and October 2023. It focuses on the prevalence of ESBL-producing Enterobacterales and related antimicrobial resistance patterns in Singapore, covering both hospital and community settings. The data, extracted from peer-reviewed articles and published abstracts, includes information on pathogen types and antibiotic susceptibility testing results. It encompasses studies from human, animal, and environmental sectors, providing a comprehensive overview of the AMR landscape in Singapore. The prevalence of resistance was pooled toward specific antibiotics in each sector. Experts are invited to respond to one of the following sections bases on their specific area of expertise:

- **Part A:** Hospital Setting
- **Part B:** Animal/Food Sector

Please provide the rationale behind each of your answers after making your estimations.

A. Human sector

| 1. In Singapore's hospitals and community healthcare settings from 2015 to 2020, what percentage of Enterobacterales isolates do you estimate were non-susceptible to 3rd generation cephalosporins but susceptible to carbapenems? | | | | |
| --- | --- | --- | --- | --- |
| 5% | 25% | 50% | 75% | 95% |
| 2. What is your estimate of the resistance rate to Amikacin among Enterobacterales isolates from patients in Singaporean hospitals and community clinics between 2015 and 2020? | | | | |
| 5% | 25% | 50% | 75% | 95% |
| 3. In the period 2015-2020, what percentage of Enterobacterales isolates from human patients in Singapore's healthcare facilities do you believe were resistant to Amoxicillin/clavulanic acid? | | | | |
| 5% | 25% | 50% | 75% | 95% |
| 4. Estimate the resistance rate to Aztreonam among Enterobacterales isolates collected from human patients in Singaporean hospitals and community health centers from 2015 to 2020. | | | | |
| 5% | 25% | 50% | 75% | 95% |
| 5. What proportion of Enterobacterales isolates from human clinical samples in Singapore's healthcare system do you think were resistant to Carbapenems during the 2015-2020 period? | | | | |
| 5% | 25% | 50% | 75% | 95% |
| 6. Estimate the percentage of Enterobacterales isolates resistant to Ceftriaxone among samples collected from patients in Singaporean hospitals and community clinics between 2015 and 2020. | | | | |
| 5% | 25% | 50% | 75% | 95% |
| 7. What is your estimate of the resistance rate to Ciprofloxacin among Enterobacterales isolates from human patients in Singapore's healthcare settings from 2015 to 2020? | | | | |
| 5% | 25% | 50% | 75% | 95% |
| 8. What percentage of Enterobacterales isolates from human clinical samples in Singaporean hospitals and community healthcare facilities do you believe were resistant to Gentamicin during the 2015-2020 period? | | | | |
| 5% | 25% | 50% | 75% | 95% |
| 9. Estimate the resistance rate to Meropenem among Enterobacterales isolates collected from human patients in Singapore's healthcare system between 2015 and 2020. | | | | |
| 5% | 25% | 50% | 75% | 95% |
| 10. What proportion of Enterobacterales isolates from human clinical samples in Singaporean hospitals and community health centers do you think were resistant to Sulfamethoxazole/Trimethoprim from 2015 to 2020? | | | | |
| 5% | 25% | 50% | 75% | 95% |

B. Animal/food sector

| 1. What was the prevalence of ampicillin-resistant *E. coli* isolates in animal samples from Singapore between 2014-2017? | | | | |
| --- | --- | --- | --- | --- |
| 5% | 25% | 50% | 75% | 95% |
| 2. Estimate the prevalence of ESBL-producing E. coli in food samples collected in Singapore between 2017-2018? | | | | |
| 5% | 25% | 50% | 75% | 95% |
| 3. What was the prevalence of tetracycline-resistant *E. coli* isolates in animal samples from Singapore between 2014-2017? | | | | |
| 5% | 25% | 50% | 75% | 95% |
| 4. Estimate the prevalence of *E. coli* resistant to amoxicillin/clavulanic acid in animal samples collected in Singapore from 2014 to 2017. | | | | |
| 5% | 25% | 50% | 75% | 95% |
| 5. What was the prevalence of nalidixic acid-resistant *E. coli* in animal samples from Singapore during 2014-2017? | | | | |
| 5% | 25% | 50% | 75% | 95% |
| 6. Estimate the prevalence of chloramphenicol-resistant *E. coli* isolates in animal samples collected in Singapore between 2014-2017. | | | | |
| 5% | 25% | 50% | 75% | 95% |
| 7. What was the prevalence of ciprofloxacin-resistant *E. coli* in animal samples from Singapore during 2014-2017? | | | | |
| 5% | 25% | 50% | 75% | 95% |
| 8. Estimate the prevalence of E. coli resistant to Gentamicin in animal samples collected in Singapore in 2017. | | | | |
| 5% | 25% | 50% | 75% | 95% |
| 9. Estimate the prevalence of E. coli resistant to Meropenem in animal samples collected in Singapore in 2017. | | | | |
| 5% | 25% | 50% | 75% | 95% |
| 10. Estimate the prevalence of ceftriaxone-resistant *E. coli* in animal samples collected in Singapore during the 2014-2017 period. | | | | |
| 5% | 25% | 50% | 75% | 95% |

C. **Variable of interest**

These are the primary focus of the elicitation and are questions that cannot be adequately addressed with existing data or models, necessitating expert judgment. In our study, the variables of interest include:

- Prevalence of ESBL Enterobacterales sampled from reservoir: This variable aims to estimate the proportion of samples from the reservoir (e.g., animals, water sources) with ESBL-producing Enterobacterales.
- Reservoir-Human Contact Rate: This variable seeks to quantify the frequency of contact between human populations and reservoirs (e.g., animals, water sources) that may harbor ESBL-producing Enterobacterales.
- Probability of Successful Transmission for Each Contact: This variable estimates the likelihood that a single contact between a reservoir (e.g., animals, water sources) and a human will result in the transmission of ESBL-producing Enterobacterales.

Experts should answer either hospital setting (part A) or community setting (part B and C,) based on their specific area of expertise. These variables are crucial for understanding the dynamics of ESBL Enterobacterales transmission within community settings and developing effective intervention strategies. Please provide the rationale behind each of your answers after making your estimations.

A. Hospital Environment:

| a) In a typical 24-hour period in 2024, how many times does an average patient in a Singaporean public hospital medical ward make skin contact with high-touch surfaces (e.g., bed rails, call buttons, tray tables) within arm's reach of their bed? | | | | |
| --- | --- | --- | --- | --- |
| 5% | 25% | 50% | 75% | 95% |
| b) In a single routine environmental sampling of 100 cm² areas on 100 different high-touch surfaces in Singaporean public hospital medical wards in 2024, how many samples would you expect to test positive for ESBL-producing Enterobacterales? | | | | |
| 5% | 25% | 50% | 75% | 95% |
| c) Consider a scenario in a Singaporean public hospital in 2024, if a patient makes skin contact with a surface contaminated with ESBL-producing Enterobacterales, what is the chance* that this single contact will result in colonization of the patient?   *Please express your answer as a number between 0 and 1, where 0 means no chance of colonization and 1 means certain colonization. For example, 0.001 would represent a 1 in 1000 chance. | | | | |
| 5% | 25% | 50% | 75% | 95% |

B. Companion Animals:

| a) What percentage of household companion animals (dog and cat) in Singapore would you expect to test positive for ESBL-producing Enterobacterales in fecal samples in 2024? | | | | |
| --- | --- | --- | --- | --- |
| 5% | 25% | 50% | 75% | 95% |
| b) For companion animals (dog and cat)-owning households in Singapore in 2024, how many times per day on average does a household member have direct physical contact (petting, playing, grooming) with their dog or cat? | | | | |
| 5% | 25% | 50% | 75% | 95% |
| c) Consider a scenario in Singapore in 2024, if an individual has direct contact with an ESBL-positive household companion animals, what is the chance* that this single contact will result in colonization?   *Please express your answer as a number between 0 and 1, where 0 means no chance of colonization and 1 means certain colonization. For example, 0.001 would represent a 1 in 1000 chance. | | | | |
| 5% | 25% | 50% | 75% | 95% |

C. Community environment:

| a) On an average weekday in 2024, how many times does a typical working adult in Singapore make skin contact with shared surfaces (e.g., door handles, handrails, touchscreens) in public spaces during their commute and work hours? | | | | |
| --- | --- | --- | --- | --- |
| 5% | 25% | 50% | 75% | 95% |
| b) In a 2024 environmental sampling study of high-touch surfaces in Singapore's public transportation system (MRT and buses), researchers collect 50 samples. How many of these samples would you expect to test positive for ESBL-producing Enterobacterales? | | | | |
| 5% | 25% | 50% | 75% | 95% |
| c) Consider a scenario when in a public space in Singapore in 2024, if an individual makes skin contact with a surface contaminated with ESBL-producing Enterobacterales, what is the chance * that this single contact will result in colonization?   *Please express your answer as a number between 0 and 1, where 0 means no chance of colonization and 1 means certain colonization. For example, 0.001 would represent a 1 in 1000 chance. | | | | |
| 5% | 25% | 50% | 75% | 95% |

**Appendix 2: Data for seed questions**

| **Sector** | **Antibiotics** | **Pooled Prevalence (95% CI)** | **Number of resistances** | **Total isolates tested** | **Number of studies** | ***I^2^*** | **Seed question ID** |
| --- | --- | --- | --- | --- | --- | --- | --- |
| animal | Ampicillin | 54.7% (18.8-88.0%) | 26 | 46 | 2 | 0.86 | C1 |
| food | ESBLs | 15.8% (2.1-39.0%) | 73 | 421 | 2 | 0.97 | C2 |
| animal | Tetracycline | 24.7% (8.4-46.1%) | 12 | 46 | 2 | 0.59 | C3 |
| animal | Amoxicillin/clavulanic acid | 18.1% (4.8-37.5%) | 9 | 46 | 2 | 0.56 | C4 |
| animal | Nalidixic acid | 14.9% (6.2-26.5%) | 7 | 46 | 2 | 0.00 | C5 |
| animal | Chloramphenicol | 11.5% (1.5-29.1%) | 6 | 46 | 2 | 0.58 | C6 |
| animal | Ciprofloxacin | 8.4% (2.2-18.0%) | 4 | 46 | 2 | 0.00 | C7 |
| animal | Gentamicin | 2.1% (1.7-16.7%) | 2 | 46 | 2 | 0.72 | C8 |
| animal | Meropenem | 0.0% (2.1-2.1%) | 0 | 46 | 2 | 0.00 | C9 |
| animal | Ceftriaxone | 1.1% (0.8-8.7%) | 1 | 46 | 2 | 0.43 | C10 |
| human | 3GC non-susceptible, carbapenem susceptible | 42.4% (32.9-52.2%) | 42 | 99 | 2 | 0.00 | A1 |
| human | Amikacin | 2.2% (1.5-3.1%) | 28 | 1265 | 2 | 0.00 | A2 |
| human | Amoxicillin/clavulanic acid | 13.6% (1.0-37.0%) | 368 | 1511 | 3 | 0.99 | A3 |
| human | Aztreonam | 1.2% (0.2-3.0%) | 3 | 246 | 1 | NA | A4 |
| human | Carbapenem | 6.8% (2.0-14.0%) | 7 | 99 | 2 | 0.34 | A5 |
| human | Ceftriaxone | 30.3% (27.8-32.8%) | 383 | 1265 | 2 | 0.00 | A6 |
| human | Ciprofloxacin | 28.2% (13.7-45.6%) | 443 | 1265 | 2 | 0.84 | A7 |
| human | Gentamicin | 4.7% (0.0-18.3%) | 157 | 1511 | 3 | 0.98 | A8 |
| human | Meropenem | 2.0% (1.3-2.8%) | 25 | 1265 | 2 | 0.00 | A9 |
| human | Sulfamethoxazole/trimethoprim | 24.2% (13.8-36.4%) | 359 | 1265 | 2 | 0.72 | A10 |
